# Nickel and Dimed: How a Common Earth Element is Short-Changing Our Health

**DOI:** 10.64898/2026.06.05.26355010

**Authors:** Chris Heitzig, David Rehkopf

**Author notes:** Corresponding author: Chris Heitzig. This study was approved by the Stanford University Institutional Review Board (Protocol No. 51310), which granted a waiver of informed consent and HIPAA authorization for the use of retrospective electronic health record data. The authors declare no competing interests. Neither the authors nor their institutions received payments or services in the past 36 months from a third party that could be perceived to influence, or give the appearance of potentially influencing, the submitted work. Funding: This work received no funding. Neither the authors nor their institutions received payment or services from a third party for any aspect of the submitted work.

## Abstract

Nickel has been studied for a long time as an environmental contaminant but less so in its connection to population health. It does not announce itself as loudly as its transition metal brethren like mercury and cadmium, but its chemical properties permit it to be deleterious as a low-dose, chronic exposure, particularly among those with immune systems sensitized to it. There is a growing evidence base and vocabulary to discuss nickel’s effect on health. However, in the U.S., there are not recent, reliable estimates of the share of the population with a nickel allergy, let alone how much nickel Americans are exposed to through their diet. This paper seeks to close this evidence gap by creating a new dataset of dietary nickel and other heavy metal exposure and assessing how high levels of dietary nickel exposure shape local demand for health care services. We use soil data from the U.S. Geological Survey and data on agricultural product transport from FoodFlows.org to create a county-level dietary nickel exposure index. We then use a large electronic health record database and double machine learning to estimate how demand for primary care services varies across levels of dietary nickel exposure. We find that counties with high nickel exposure experience an increase in the share of primary care office visits for symptoms highly suggestive of nickel poisoning. This result survives multiple hypothesis test corrections and placebo tests. Our research suggests that nickel has harmful effects on individual health whose exposure can be measured at a population level, and is shaping primary care across the U.S.

## 1. Introduction

The food we eat matters, but so does where it is grown. Industrialization and commercial farming practices have introduced heavy metal contamination into the soil that produces the food we eat. But much of the variation in heavy metal content in soil reflects geologic sources that are millions of years in the making (1). Species of crops are now grown in very different environments in which they evolved (2). For some of these crops, accumulating heavy metals was evolutionarily beneficial for flourishing, and they “learned” to take what scarce metals were available in the soil.

The evolutionary mismatch between where crops evolved and where they now live poses issues for the health of Americans. Crops grown in parts of the U.S. where soil heavy metal content is high may have high levels of those heavy metals. The negative health effects of certain heavy metals like mercury are well publicized, but not as much research has focused on nickel and its affects on the health of those ingesting it.

The properties of nickel make it highly problematic and a likely target by the immune system (3). Its electron structure means that it binds readily to proteins throughout the body. Moreover, because it occurs in high concentrations in objects like jewelry with which we come into frequent contact, people are disposed to developing allergies to it (4). If the immune system in sensitized individuals encounters protein-nickel complexes, it may mount an immune response, resulting in symptoms like dermatitis, elevated heart rate or palpitations, head aches, anxiety, cough, inflammation, and general malaise, a condition known as systemic nickel allergy syndrome (SNAS) (3, 5, 6). It is difficult to say how many Americans have a nickel allergy, but estimates from Europe suggest that the figure may be between 10 and 20 percent of the population (4, 7). Because nickel is neither an essential nutrient nor potently poisonous, it has existed in the shadow of other minerals (the United States Department of Agriculture, for example, does not collect data on nickel) (8, 9). For this and other reasons, we do not have a good sense of how much nickel Americans are exposed to in their diet and what effect this may have on their health.

This paper seeks to address this gap by answering the question “Does the level of nickel in the food people consume correlate with health conditions consistent with an immune-mediated response to nickel compounds?” To answer this question, we build a dietary nickel exposure index by combinding two different data sources: 1) Soil samples from the U.S. geological survey and 2) Data from FoodFlows.org that maps the county food is grown in to the county in which it is consumed. The result is a county-level estimate of the exposure to dietary nickel that stems from the soil in which it is grown. We then build an econometric model that utilizes double-machine learning to estimate how dietary nickel exposure shapes health care utilization, for which we use a large, nationally-representative electronic health records database. We are able to see whether people in high-nickel counties visit their primary care clinic for different reasons than those in low-nickel counties, and whether this is consistent with the symptoms predicted by high serum nickel levels.

Because the effects of nickel exposure in sensitized individuals are immune-mediated, we also test the hypothesis that nickel exposure plays a role in the development of autoimmune conditions. More specifically, we compare the test positivity rate of two relatively common antibody tests for Hashimoto’s thyroidititis (thyroid peroxidase) and type 1 diabetes (beta cell antibodies). The chemical properties of the former make it a likely host for interactions with nickel, whereas the opposite is true of the latter.

First, we find that higher levels of dietary nickel are associated with an increase in primary care office visits for reasons consistent with what is predicted by biology: heart palpitations, migraines, panic attacks, and cough. Moreover, when we run a placebo test against ten conditions that biology would suggest to have no relation to dietary nickel exposure, the results largely show no association. Second, after correcting for multiple hypothesis testing, the strongest result that emerges is the effect on heart palpitations. We note that the relationship is nonlinear: the effect on visit share becomes more pronounced at higher levels of nickel exposure. Finally, the tests on antibody positivity do not reveal a clear relationship between nickel and antibody prevalence. However, the test is also fairly underpowered, and the results suggest a deeper investigation into whether dietary nickel exposure drives certain autoimmune conditions.

## 2. Biological and clinical framework

It is believed that a contact allergy to nickel is a prerequisite for a symptomatic reaction to nickel in the bloodstream, although this is not proven. The exact share of Americans with a nickel allergy is not known, since a vast majority of screening comes from the population referred to allergy clinics. The last reliable estimate of population level prevalence in the U.S. came in 1979 and was 5.8 percent (7). Estimates from Europe generally place the population incidence between eight and 19 percent (4). The authors have not found a reliable estimate of what share of the population have a proven systematic reaction to ingested nickel, a condition known as systemic nickel allergy syndrome or SNAS.

While incidence of SNAS is not known, nickel allergies are shockingly co-morbid with other immune-modulating conditions. A study of 239 patients referred to the Department of Immune-Mediated Inflammatory Diseases of the Padre Pio Hospital in Italy found that 57 percent of these patients had SNAS, the most common condition they for which they tested (10). This group also had a higher incidence of autoimmune thyroiditis. Moreover, a study of endometriosis patients revealed that more than 90 percent of them had a nickel allergy. While these studies do not show that nickel plays a causal role in the development of these conditions and others, it suggests that nickel plays some role in the disease process, either as a driver, a co-morbidity, or as an outcome.

In the United States, nickel is regularly included in alloys that make jewelry. Nickel has been shown to be a highly immunogenic element, for a variety of reasons. Nickel is a transition metal that readily loses two electrons and forms new bonds. When sweat comes into contact with nickel-containing jewelry, nickel ions break off and are carried into the skin. There, they bond to skin proteins and form nickel-protein complexes, that together form a new antigen. Immune cells present these complexes to T cells, and, in some people, the immune system becomes sensitized, or allergic,^*^ to nickel.

The immune system that reacts in the skin is part of the same immune system that circulates throughout the body. That means that, among sensitized individuals, it is possible that serum nickel levels incite an immune response even though the encounter occurs in a different part of the body. In vitro tests show that encounters with these nickel-protein complexes, or “haptens”, produce a broad immune response: 21 of 33 immune molecules tested were elevated. Changes in these molecules are linked with symptoms like fatigue, brain fog, malaise, increased sympathetic nervous system dominance, aches, pain, and headache (11). Moreover, these molecules can cross the blood brain barrier and alter signaling in the brain, resulting in anxiety, depression, and intrustive thoughts (11).

What makes nickel so potent as an environmental pollutant is its combination of ubiquity and immunogenicity. While it is not the only immunogenic metal (there is a reason aluminum salts are included in vaccines), it is strongly immunogenic. It is found in abundance in the soil, in the water we drink, and in the built environment we occupy. It is found in oral metal alloys used in braces, retainers, and tooth fillings, creating a persistent low-dose exposure that excites the immune system of susceptible individuals.

### A. Dietary nickel heterogeneity across foods and places

Nickel predominantly enters the body through the food and liquids humans ingest. What made SNAS so difficult to discover was that the concentration of nickel not only varies by type of food but also by the environment in which it is grown. Some plants utilize much more nickel than other plants, and thus, if present, it accumulates in the plant (8, 12). High nickel accumulators include tree nuts, legumes, and whole grains. The amount of nickel in a given plant, however, depends on soil nickel bioavailability (concentration, pH level, presence of other chemicals). Legumes grown from one location may cause severe contact dermatitis in a patient, while legumes from another location may cause no noticeable symptoms at all.

The USDA does not collect measurements on nickel present in the food, so it is difficult to say how nickel is present in foods. Data from Europe, most notably from the European Food Safety Authority (ESFA), show that nickel is highest among foods like whole grains, nuts, seeds, legumes, and beans (12, 13). Lentils, for instance, contain an average of 151mcg of nickel per 80 grams. Eggplant, by comparison, contains 1.4mcg of nickel per 80 grams. The variability of nickel within foods is more striking than the variability across foods. The samples taken by ESFA showed that lentils contained anywhere from 58.7 to 296 mcg of nickel per 80 grams, more than a fivefold change. The range is the difference between moderate consumption without symptoms and a flare up in sensitive individuals.

The USDA estimates that around 84 percent of food consumed by Americans is domestically sourced, with the remaining 16 percent imported from other countries (14).

## 3. Data and measurement

### A. Outcomes

The core outcome variable—share of outpatient doctor’s visits in a county—is sourced from the American Family Cohort (AFC), a large electronic health record (EHR) focused on primary care. AFC consists of approximately 90 million office visits from eight million patients and covers all 50 states (15). We build our cohort by first considering only the years after 2012, because it is the first year that our exposure measure is available (more on this below). We consider data over a ten-year period through 2022. Furthermore, we keep data from primary care cites that report at least, on average, one visit per day for every year of the study period 2012-2022. This ensures that the outcomes are not altered simply by the partial availability of AFC data. Because our analysis is spatial, we drop those office visits that do not report a county where the patient lives. It is important to use the county in which a patient lives rather than the county of the provider, as the exposure occurs in the neighborhood in which the patient lives.

We use phecodes to coarsen International Classification of Diseases 10th revision (ICD-10) codes (16). We drop any codes that do not have a corresponding match to a phecode and aggregate to the county level. Any phecode-county buckets that have fewer than 25 observations are categorized as “Other”.

Our preferred outcome measure is the share of office visits in a given county for a given condition over the period 2012-2022. The reason is that we want to know if nickel exposure changes the reason people seek care, not whether nickel exposure changes the overall healthcare burden.^†^ Let *V*_*dc*_ denote the number of outpatient office visits in county *d* assigned to condition *c* over 2012–2022, and let 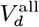 denote total outpatient office visits in county *d* over the same period. We construct the county-level outcome share as

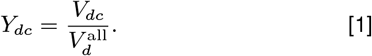

One condition that our biological framework and the previous literature suggest might be affected by nickel exposure is thyroid autoimmunity. The phecodes do not allow us to separate thyroid autoimmunity from other thyroid issues such as non-autoimmune hypothyroidism and hyperthyroidism, thyroid cancer, and goiter. To shed light on the potential mechanism behind these visits, we instead use the test positivity rate for thyroid-related antibodies. As a comparison, we use beta cell antibodies, which do not bind well to nickel and thus make for a good comparison. We use the test positivity rate to control for how much testing occurs in a given county.

### B. Exposure

The exposure studied in this paper is an estimate of nickel content in the food. We create an index of exposure to nickel in food using soil samples from the United States Geological survey (USGS) and data from FoodFlows.org that the county of food production to county of food consumption. From 2007 to 2013, the USGS collected soil samples for 45 elements at a 40km resolution. For the FoodFlows.org data, only raw agricultural products are considered in the food flows data. In cases where there are multiple soil samples in a given county, we take a simple average of these soil samples to construct a county-level estimate of the nickel soil content. We then construct the the “food nickel index” as follows:

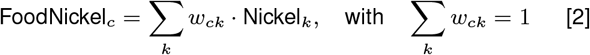

where Nickel_*k*_ denotes the average soil nickel level in county *k*, and *w*_*ck*_ represents the share of raw agricultural products in county *c* sourced from county *k*. Because Americans consume a vast majority of their diet from domestically produced food, we believe this index reflects true dietary nickel exposure.

In carrying out this research, we considered another concept of nickel exposure, namely, “Destination Soil Exposure”, that captures direct exposure to the concentration of soil nickel in the county in which a patient lives. However, we do not examine this concept in detail, as we believe that nickel is not present in sufficiently high concentrations in any soil to engender an immune response. Note that exposure to agricultural products grown in the same county a patient lives in is incorporated in the Food Nickel Index.

One limitation of the USGS soil sample data is that the publicly available data do not include raw measurements but rather the decile into which a measurement falls. For the nickel soil samples, for example, they are distributed across percentile bins as follows: the 0–10th percentile corresponds to concentrations from less than 0.5 to 4.0 mg/kg; the 10–20th percentile from 4.0 to 6.6 mg/kg; the 20–30th percentile from 6.6 to 8.9 mg/kg; the 30–40th percentile from 8.9 to 11.2 mg/kg; the 40–50th percentile from 11.2 to 13.5 mg/kg; the 50–60th percentile from 13.5 to 15.7 mg/kg; the 60– 70th percentile from 15.7 to 18.3 mg/kg; the 70–80th percentile from 18.3 to 21.6 mg/kg; the 80–90th percentile from 21.6 to 28.1 mg/kg; and the 90–100th percentile from 28.1 to 1810 mg/kg.

For this reason, *Nickel*_*k*_ represents the percentile of all soil nickel measurements across the US, and thus *FoodNickel*_*c*_ can be thought of as the percentile of nickel exposure through the domestic food system. Certainly, one’s true exposure depends also on the composition of one’s diet; however, this is beyond the purview of this paper.

### C. Covariates and controls

We use data from the American Community Survey (ACS) to control for factors within a given county and the counties from which it imports its food that might affect the reasons people utilize primary care. We use the full suite of ACS variables as controls, which cover topics such as demographics, education, income, poverty, employment, housing characteristics, and others. Some specifications use principle component analysis (PCA) to reduce the dimensions of the ACS data. These specifications use principle components for both the destination county and the origin county.

When creating PCAs from ACS characteristics in the destination county, we follow a standard process. For destination county *d*, let 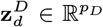 denote the stacked destination-side controls. We standardize each variable as

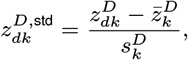

and form destination PCs from the leading eigenvectors of the sample covariance matrix,

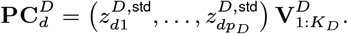

We follow a slightly different process when constructing shipment-weighted origin controls and origin PCAs. For each destination county, we take a weighted average of the ACS characteristics of the origin counties, weighted by the amount of food provided by the origin county for the destination county. We then standardized these weighted averages and create PCAs out of them. Mathematically, let *w*_*od*_ be the food-flow weight from origin *o* to destination *d*, normalized so ∑_*o*_ *w*_*od*_ = 1. For each origin-side characteristic *m*,

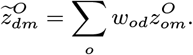

After standardizing 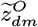 across destinations, origin PCs are

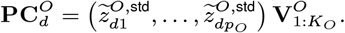

Figure 1 reports the cumulative variance explained by the leading 20 Census-based principal components in this dimension-reduction step for destination counties. In specifications that use these vectors, we limit the set of controls to the first seven components.

**Fig 1.**
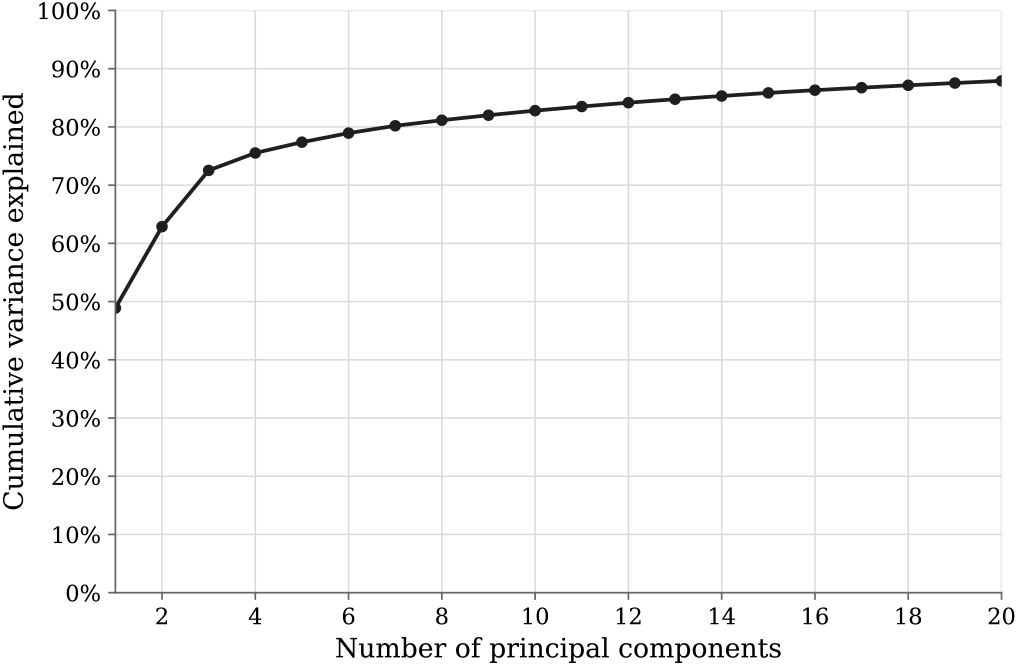
Cumulative variance explained by the leading Census PCA components. *Note*: The figure plots the cumulative share of variance explained by the top 20 principal components from the standardized Census-based county covariates used in the PCA construction step.

## 4. Empirical strategy

### A. Baseline nonlinear specification

This section outlines how we analyze the relationship between the visit share of conditions of interest and food nickel content. Our biological framework suggests that there might be a nonlinear relationship between condition share and food nickel. For this reason, we first estimate a condition-specific adaptive spline on the raw county-level outcome, controlling directly for the first seven destination PCA components, the first seven origin PCA components, and state fixed effects.^‡^ Let *Y*_*dc*_ be the visit-share outcome for condition *c* in county *d, T*_*d*_ the food nickel index, and 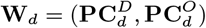 the baseline PCA control stack. The baseline adaptive-spline specification is

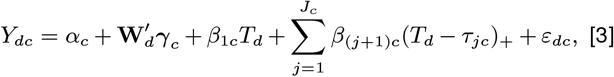

where *J*_*c*_ ∈ {0, 1, 2, 3} is chosen condition by condition, (*x*)_+_ = max {*x*, 0}, and the breakpoints 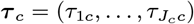 are selected from a candidate percentile grid using the BIC-selected specification. Standard errors are clustered at the state level. Figure 2 visualizes the condition-specific fits from Eq. (3).

**Fig 2.**
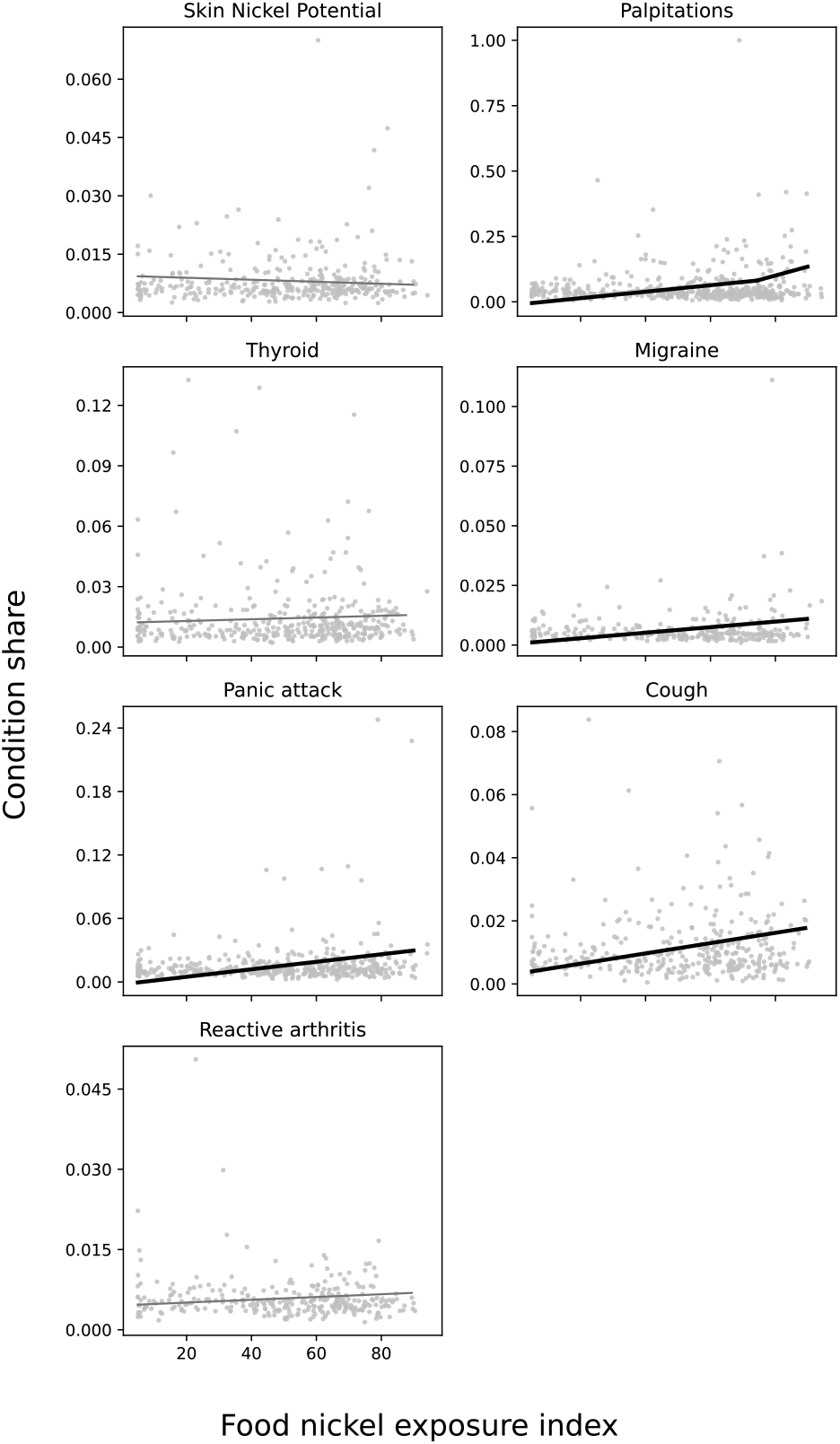
Condition-panel adaptive spline array from the baseline nonlinear specification. *Note*: Each panel reports the fitted adaptive-spline relationship from the baseline nonlinear specification in Eq. (3). The plotted points use the raw county-level outcome and exposure data, not DML-residualized values. The DML step is then run on the condition-specific spline basis selected from this stage, as in Eq. (6).

**Fig 3.**
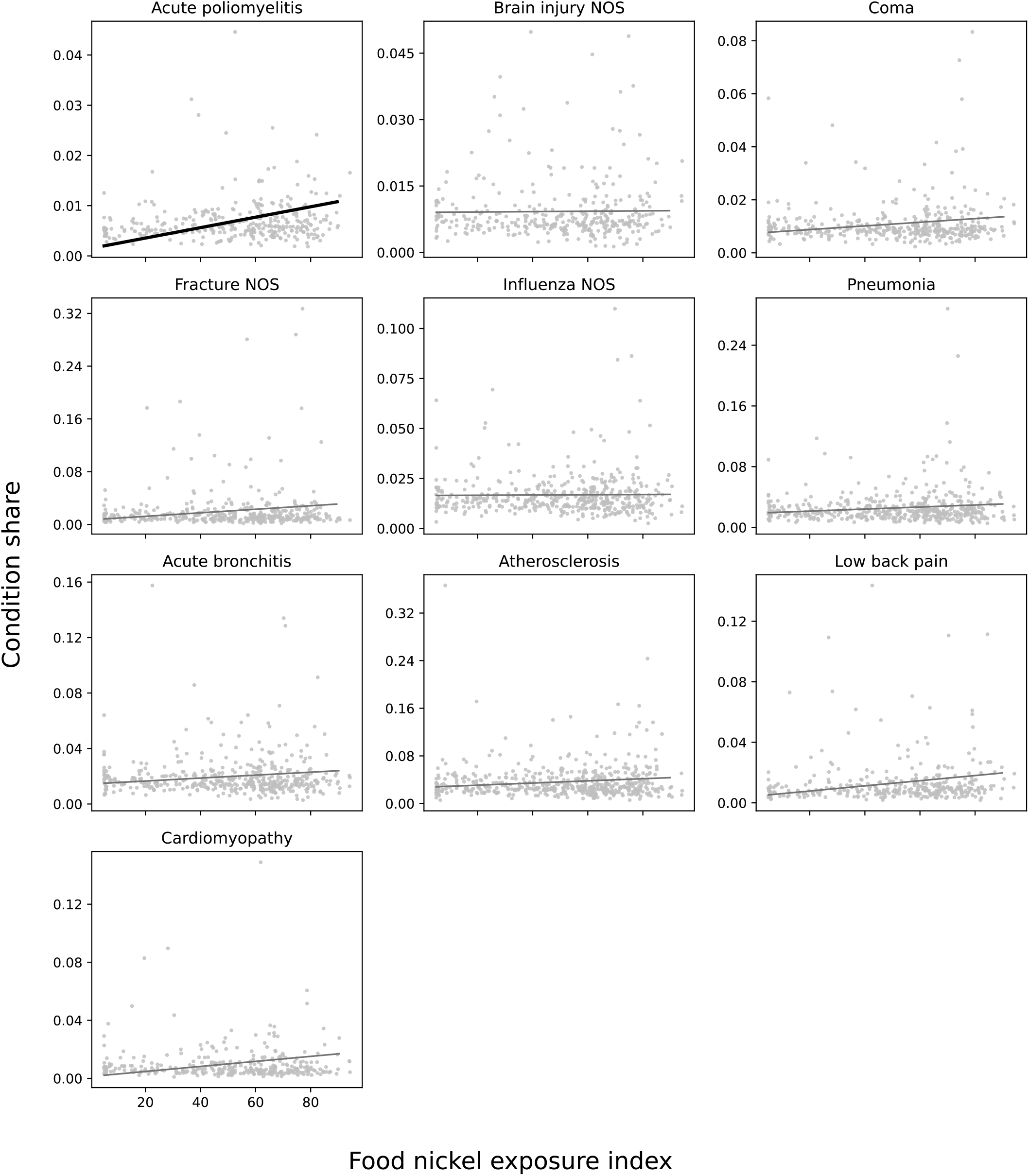
Condition-panel adaptive spline array for placebo-condition outcomes under the DML specification. *Note*: This figure applies the same adaptive-spline first-stage specification as Eq. (3) to conditions that are not theorized to be associated with SNAS and are used here as placebo outcomes. The plotted points use the raw county-level outcome and exposure data, not DML-residualized values.

### B. Residualizing confoundedness from mineral co-occurrence

One limitation of Equation 3 is that it does not account for the possibility that minerals that co-occur with nickel (e.g. cobalt) are actually driving potential health outcomes rather than nickel itself. To address this concern, our preferred specification utilizes double machine learning (DML) to residualize our outcome and exposure of variation in induced by the pretense of important groups of minerals (17, 18). We include a mineral that co-occurs often with nickel (cobalt); a chemicaly adjacent transition metal (chromium); a mineral that enters the body via the DMT1 enzyme and whose toxicity could theoretically influence similar health issues (Manganese, iron and aluminum); nutritionally necessary minerals that compete for absorption and binding (copper and zinc); minerals that have shown to be toxic to the human body (lead, cadmium, arsenic, and mercury); and minerals that could interact with nickel in the body (selenium and molybdenum); and finally other potentially confounding minerals in the environment (antimony, uranium, vanadium, and barium). We choose to estimate the adaptive spline before residualizing via DML because we want to residualize the different splines that exist, as DML does not correspond to the data generating process and is instead focused on prediction, without regard to the underlying relationship that may exist between the condition share and the food nickel exposure index. Our use of the full ACS covariate set is similar in spirit to prior high-dimensional applications using ACS data with double-selection methods, such as Bach, Chernozhukov, and Spindler’s analysis of wage gaps using the 2016 American Community Survey (19).

More formally, let *X*_*d*_ denote the full DML control stack (destination census variables and origin census variables captured by the PCAs;^§^ baseline covariates like a log of domestic food imports and a log of the total number of office visits reported); and non-nickel minerals listed above. For each condition *c*, let *B*_1*dc*_ = *T*_*d*_ and, for 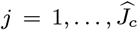, let 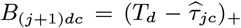 denote the spline basis implied by the model selected in Eq. (3). Let 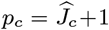 be the number of basis terms in that selected spline. In each cross-fitting fold *k*, nuisance functions are estimated using ElasticNet regularization as

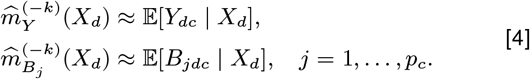

Orthogonalized variables are then

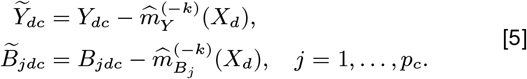

The DML second stage is estimated on this fixed, condition-specific spline basis:

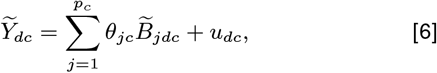

so the adaptive-spline search is completed first and is not rerun after residualization. For the one-break case, define 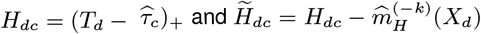; the estimating equation is

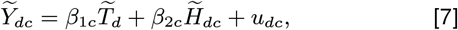

with 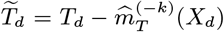 and state-clustered standard errors. We also adjust p-values for multiple hypothesis testing using Benjamani-Hochberg FDR correction.

### C. Placebo and robustness analyses

As a robustness check, we check whether our main results are not spuriously significant artifacts of the data by testing a number of placebo conditions which previous research would suggest have no theoretical relation to SNAS. These include acute poliomyelitis, brain injuries, coma, fractures, influenza, pneumonia, acute bronchitis, atherosclerosis, low back pain, and cardiomyopathy.

### D. Tissue-affinity contrast strategy

Our biological framework suggests that tissues in the body with greater binding affinity to nickel may be casualties of the immune system as it targets nickel-laden compounds. As part of its effort to eradicate these compounds, the immune system could develop antibodies to the tissues to which nickel binds. In Section 5 we test whether this is true by comparing the test positivity rate of two commonly examined antibodies: Thyroid peroxidase antibodies (TPOAb) and beta cell antibodies (AAb).

For this purpose, we utilize Equation 6. Because of its relative affinity for nickel, we suspect that the coefficient on TPOAb may be positive. We examine AAb as a placebo test, as its properties do not permit it to bind readily to nickel. We therefore suspect that there will be no association between the AAb test positivity rate and the food nickel exposure index.

We source our data on these tests from the AFC database. One limitation, however, is that, while these tests are common, only 192 counties report at least 30 TPOAb tests from 2012-2022, while only 40 do so for beta cell antibodies. We therefore only have the statistical power to detect very large effects.

## 5. Results

### A. Spline break identification

Figure 4 displays the results of Equation 3. It shows the raw association between outcome visit share and the food nickel exposure index, controlling for destination and origin PCAs and state fixed effects, for the 53 conditions for which we have data. Relationships that are statistically significant at the 10 percent level are in bold. Including the seven spline breaks identified, there are 60 relationships tested in total, of which 13 are statistically significant. These include many of the conditions that we hypothesized in Section 2. Equally important, none of the statistically significant slopes are negative; if these findings were driven primarily by spurious statistical significance, we would expect some significant slopes to point in the opposite direction. This directional asymmetry suggests a systematic signal rather than a collection of random false positives.

**Fig 4.**
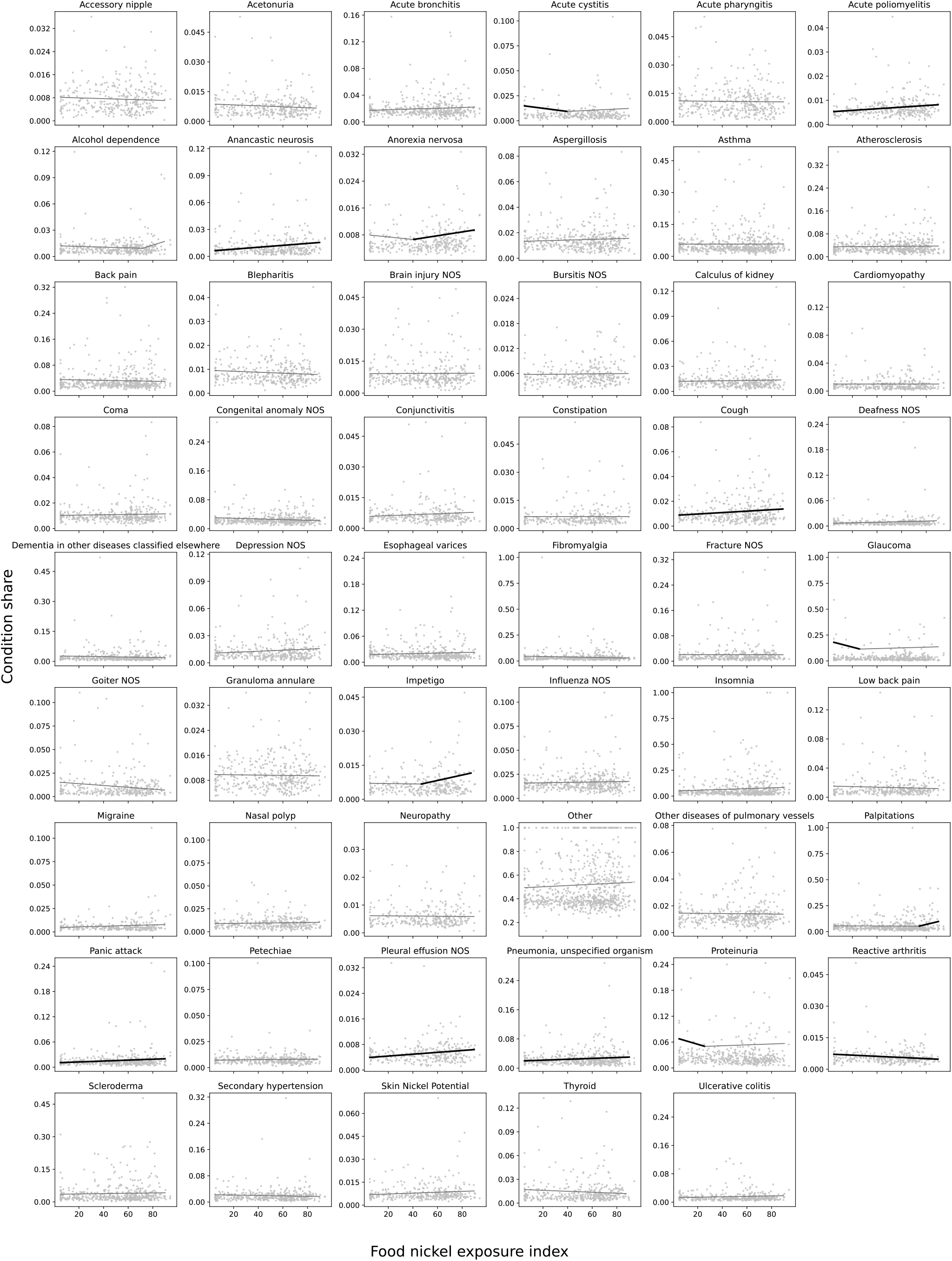
Condition-panel adaptive spline array for all conditions with state fixed effects and PCA controls. *Notes*: Each panel plots county-level condition shares against the food nickel exposure index. Gray points show the raw county observations. Solid fitted lines report adaptive spline estimates using the same piecewise-linear spline form as Eq. (3), here estimated in condition-specific regressions that control for state fixed effects, destination PCA components 1-7, and origin PCA components 1-7. Knot locations are chosen condition by condition from candidate percentiles using the BIC-selected specification. Line segments are shown in bold when the corresponding segment slope has a p-value below 0.10. “Skin nickel potential” refer to a group of skin conditions plausibly related to a nickel allergy that includes allergic dermatitis, eczema, and hives.

### B. Main specification

Figure 2 displays the condition-specific fits from the DML specification with the raw county-level data plotted in the background. Of the seven conditions we identified as most likely to be affected by nickel exposure, four of them had a positive and statistically significant association with nickel exposure at the ten percent level: Heart palpitations, migraine, panic attack, and cough. In addition, two more conditions—thyroid-related issues and reactive arthritis—have a positive but statistically insignificant relationship. “Skin nickel potential” had a negative and statistically insignificant association. The second spline of palpitations had a slope beginning at the 80th percentile of the food nickel exposure index that was steeper and more statistically significant than the slope running from bottom 80 percent. The details of these estimations are contained in Table 2.

The coefficient on the four statistically significant conditions are of modest but meaningful magnitudes, except for heart palpitations, which has rather large coefficients. Table 2 shows that first spline has a slope of 0.0012, implying a 10 percentage point increase in the food nickel exposure index is associated with a 1.2 percentage point increase in the share of office visits due to heart palpitations. Above the 80th percentile of the food nickel exposure index results in more dramatic increases, whereby a 10 percent increase in the index corresponds to a 3.4 percentage point increase in the frequency of such visits.

### C. Adjusting for multiple hypothesis testing

Table 2 provides FDR-adjusted p-values using the Benjamani-Hochberg method. We apply the adjustment to all p-values from all splines in the seven conditions of interest, which in our case is eight p-values. The adjustment moves spline 1 of palpitations, migraine, panic attack, and cough to 0.109. The FDR-adjusted p-value for spline 2 of palpitations remains at 0.0244.

### D. Tissue-affinity contrast strategy

Table 1 contains the tests of the relationship between the food nickel exposure index and antibody prevalence. Both antibodies have a statistically insignificant relationship with the food nickel exposure index. The coefficient for beta cell antibodies is negative and small in magnitude, while the one for TPO antibodies is positive and fairly large. The sample size for these tests is considerably smaller than it was for those tests which used office visit share as the outcome. As a result, the statistical power is diminished for these tests. The TPO antibody tests has a p-value of 0.17, closer to significance than the beta cell antibodies by still insignificance.

**Table 1.** Antibody results.

| Outcome | N | Optimal Breaks | Segment 1 Slope | Segment 1 p-value |
| --- | --- | --- | --- | --- |
| Beta cell Ab | 40 | 0 | -0.000124 | 0.813 |
| TPO Ab | 192 | 0 | 0.000888 | 0.172 |

*Notes:* Estimates follow the same analysis procedure as the main condition-level results: (i) select adaptive spline structure for the nickel index, (ii) residualize outcome and treatment with the DML nuisance model and evaluate spline basis terms in the residualized space.

## 6. Discussion

We find that living in a county with high exposure to dietary nickel via the source of their food is associated with a higher incidence of health conditions which biology predicts is associated with an immune response to nickel. In particular, we find that people in more exposed counties seek care disproportionally more often for conditions like heart palpitations, migraines, panic attacks, and coughs—in short, an immune driven shift into a highly sympathetic state. These results persist even after accounting for destination-specific characteristics, origin-specific characteristics, mineral co-occurrence, and other factors. Among our results, the evidence is strongest for heart palpitations, whose statistical significance is preserved at the five percent level even after adjusting for multiple hypothesis testing.

At present, SNAS is a condition characterized above else by dermatitis in response to ingested nickel. This study provides evidence consistent with the idea that our current conception of SNAS is too narrow and only captures one segment of SNAS patients (i.e. those whose symptoms present most visibly).

The null result for “skin nickel potential” points in the same direction. If SNAS were best characterized primarily as dermatitis, we would expect the skin-oriented outcome to be among the strongest signals. Instead, its lack of statistical significance is consistent with the possibility that dermatitis is too nonspecific an endpoint for detecting SNAS in population-level utilization data: common causes of dermatitis may drown out the smaller signal attributable to nickel-sensitive patients.

We hypothesized that different tissues might have different affinity for nickel, and therefore that they might have different likelihoods of being involved as a casualty of the immune system responding to haptens that involve these tissues. The results in Table 1 do not necessarily support this hypothesis, but also do not contradict it. It would be useful to test this hypothesis in a setting with more statistical power. While the direction and the relatively low p-value may be tempting to conclude that more statistical power may plausibly reveal a statistically significant relationship, the fact that the coefficient on the thyroid office visit share is clearly insignificant in Table 2 suggests that reality is complex and multi-factorial.

**Table 2.**
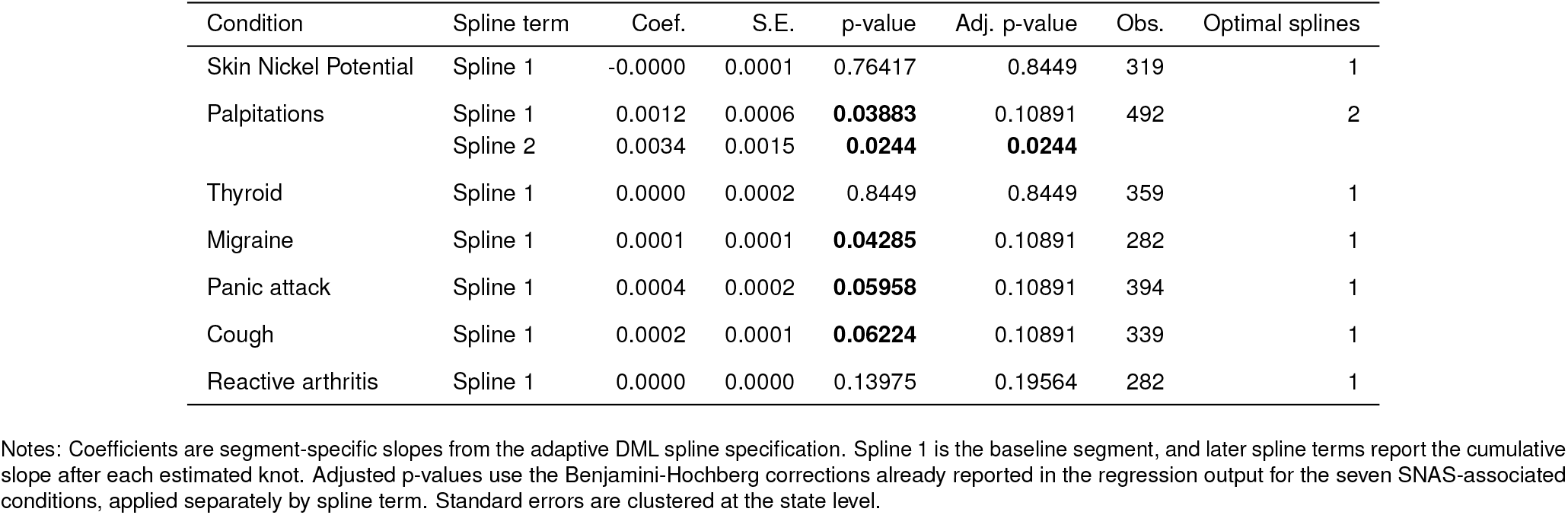
Adaptive DML spline estimates for seven SNAS-associated conditions.

| Condition | Spline term | Coef. | S.E. | p-value | Adj. p-value | Obs. | Optimal splines |
| --- | --- | --- | --- | --- | --- | --- | --- |
| Skin Nickel Potential | Spline 1 | -0.0000 | 0.0001 | 0.76417 | 0.8449 | 319 | 1 |
| Palpitations | Spline 1 | 0.0012 | 0.0006 | **0.03883** | 0.10891 | 492 | 2 |
|  | Spline 2 | 0.0034 | 0.0015 | **0.0244** | **0.0244** |  |  |
| Thyroid | Spline 1 | 0.0000 | 0.0002 | 0.8449 | 0.8449 | 359 | 1 |
| Migraine | Spline 1 | 0.0001 | 0.0001 | **0.04285** | 0.10891 | 282 | 1 |
| Panic attack | Spline 1 | 0.0004 | 0.0002 | **0.05958** | 0.10891 | 394 | 1 |
| Cough | Spline 1 | 0.0002 | 0.0001 | **0.06224** | 0.10891 | 339 | 1 |
| Reactive arthritis | Spline 1 | 0.0000 | 0.0000 | 0.13975 | 0.19564 | 282 | 1 |

Notes: Coefficients are segment-specific slopes from the adaptive DML spline specification. Spline 1 is the baseline segment, and later spline terms report the cumulative slope after each estimated knot. Adjusted p-values use the Benjamini-Hochberg corrections already reported in the regression output for the seven SNAS-associated conditions, applied separately by spline term. Standard errors are clustered at the state level.

Future work in this area should seek to expand the research at both macro and micro levels. At a macro level, such work could create an exposure index that covers all countries. It also could analyze whether the conclusions reached in this paper also hold in other countries. At a micro level, there is need to understand the biological pathways that cause SNAS-related symptoms and firmly establish whether a nickel allergy is a sufficient condition for observing these symptoms, a necessary condition, or both.

Prior work has established the immune response of sensitized patients *in vitro* (20), but future work could consider *in vivo* analysis. An ideal format for such a study is a crossover study involving sensitized (ideally a sample containing some patients with symptoms suggestive of SNAS) individuals and controls. After a one week low-dietary-nickel washout period for all participants, the sample would be divided into four treatment arms:

- Arm 1: no contact allergy, low-nickel diet.
- Arm 2: no contact allergy, high-nickel diet.
- Arm 3: contact allergy, low-nickel diet.
- Arm 4: contact allergy, high-nickel diet.

Participants would follow their diet for two weeks, take measurements, undergo a low-nickel washout diet of one week, follow the alternative treatment regime for two weeks, and finally take additional measurements. The core outcomes would be symptoms, including visual evaluations for contact dermititis, and extensive immune-marker panel, similar to those taken in de Graaf et al. (2023) (20). A comparison of the control group’s low nickel vs high nickel diet should explain whether it is possible to have an immune response to nickel even if not sensitized, while a comparison of the high nickel diet in sensitized individuals compared to a low nickel diet in controls should give a better idea of *in vivo* immune signalling in the presence of nickel haptens.

## 7. Limitations

The main drawback to this study is that, despite its many efforts to control for confounding variables, it does not utilize a source of exogenous variation and therefore limits the extent to which the results may be given a causal interpretation. The study also is limited in addressing a few sources of measurement error, particularly in that the USGS data contain percentages rather than raw measurements and that the FoodFlows.org data are estimated rather than measured.

## Data Availability

The data used in this study are from the American Family Cohort (AFC). Individual-level data cannot be publicly shared because they contain protected health information and are subject to data use agreements. Access to AFC data may be available to qualified researchers subject to approval by the data provider and relevant ethical oversight bodies.

https://med.stanford.edu/phs/data/american-family-cohort--afc-.html

## A. Annex materials

* This is a type-IV or T-cell mediated allergy, distinct from IgE mediated allergies.

† Using the raw number of office visits to explore the latter is a noisy outcome variable and prone to confounding that is difficult to control for. Such a study would benefit from larger sample sizes and a panel structure.

‡ There is no universally accepted threshold for selecting the number of principal components, and the literature commonly employs cumulative variance-explained thresholds ranging from roughly 70 percent to 90 percent. These cutoffs are inherently heuristic and context-dependent rather than theoretically derived. In our setting, seven components explained approximately 72 percent of the variation in the underlying ACS measures. While adding four additional components increased the share of explained variance by roughly eight percentage points, achieving a further eight-percentage-point increase would have required approximately thirteen additional components. We therefore selected seven components as a practical balance between dimensionality reduction and information retention, consistent with the spirit of the variance-explained heuristic.

§ We do not insert the principle components by rather the raw census data in case the variation could be repurposed for residualizing exposure and outcomes.

## REFERENCES

1 DB Smith, F Solano, LG Woodruff, WF Cannon, KJ Ellefsen, Geochemical and mineralogical maps, with interpretation, for soils of the conterminous united states, (U.S. Geological Survey), Scientific Investigations Report 2017–5118 (2019).

2 CK Khoury, et al., Origins of food crops connect countries worldwide. Proc. Royal Soc. B: Biol. Sci. 283, 20160792 (2016).

3 F Riedel, et al., Immunological mechanisms of metal allergies and the nickel-specific TCR-pMHC interface. Int. J. Environ. Res. Public Heal. 18, 10867 (2021).

4 M. Ahlström, JP Thyssen, M Wennervaldt, T Menné, JD Johansen, Nickel allergy and allergic contact dermatitis: a clinical review of immunology, epidemiology, exposure, and treatment. Contact Dermat. 81, 227–241 (2019).

5 CS Jensen, T Menné, JD Johansen, Systemic contact dermatitis after oral exposure to nickel: a review with a modified meta-analysis. Contact Dermat. 49, 227–238 (2003).

6 L Ricciardi, et al., Systemic nickel allergy syndrome: epidemiological data from four italian allergy units. Int. J. Immunopathol. Pharmacol. 27, 131–136 (2014).

7 SD Prystowsky, et al., Allergic contact hypersensitivity to nickel, neomycin, ethylenediamine, and benzocaine: relationships between age, sex, history of exposure, and reactivity to standard patch tests and use tests in a general population. Arch. Dermatol. 115, 959–962 (1979).

8 Agency for Toxic Substances and Disease Registry, Toxicological Profile for Nickel (2024).

9 U.S. Department of Agriculture, Agricultural Research Service, FoodData Central: Download data (2026).

10 M Andrioli, P Trimboli, D Maio, L Persani, M Minelli, Systemic nickel allergic syndrome as an immune-mediated disease with an increased risk for thyroid autoimmunity. Endocrine 50, 807–810 (2015).

11 R Dantzer, Cytokine, sickness behavior, and depression. Immunol. Allergy Clin. North Am. 29, 247–264 (2009).

12 EFSA Panel on Contaminants in the Food Chain (CONTAM), Update of the risk assessment of nickel in food and drinking water. EFSA J. 18, e06268 (2020).

13 AD Sharma, Low nickel diet in dermatology. Indian J. Dermatol. 58, 240 (2013).

14 U.S. Department of Agriculture, Economic Research Service, U.S. Food Imports (2025).

15 D Balraj, et al., American Family Cohort: A data resource description (2023).

16 Vanderbilt Phecode Team, Phecode Map 1.2 with ICD9 and ICD-10cm Codes (PheWAS Catalog) (2026) Accessed May 31, 2026.

17 V Chernozhukov, et al., Double/debiased machine learning for treatment and structural parameters. The Econom. J. 21, C1–C68 (2018).

18 AK Mengistu, KA Yeneakale, ND Baykemagn, ZY Melese, AE Gedefaw, Application of causal forest double machine learning (dml) approach to assess tuberculosis preventive therapy’s impact on art adherence. Sci. Reports 15, 29130 (2025).

19 P Bach, V Chernozhukov, M Spindler, Closing the u.s. gender wage gap requires understanding its heterogeneity. Oxf. Bull. Econ. Stat. 83, 380–414 (2021).

20 NPJ de Graaf, et al., Nickel allergy is associated with a broad spectrum cytokine response. Contact Dermat. 88, 10–17 (2023).

